# Altered functional connectome hierarchy with gene expression signatures in newly-diagnosed focal epilepsy

**DOI:** 10.1101/2021.07.18.21259977

**Authors:** Christophe E de Bézenac, Lorenzo Caciagli, Batil K Alonazi, Boris C Bernhardt, Anthony G Marson, Simon S Keller

## Abstract

**Objective:** Neuroimaging research is providing insights into epilepsy as a disorder of brain connectivity linked to functional impairments which may have an identifiable genetic component. This case-control study aims to identify imbalances in a functional connectome dimension spanning from unimodal to transmodal networks and explore the potential genetic basis of such alterations in patients with newly diagnosed focal epilepsy (NDfE).

**Methods:** We used gradient-based analysis of resting-sate fMRI data comparing cortical gradient maps in patients with NDfE (n = 27) to age and sex-matched controls (n = 36). Using a brain-wide gene expression dataset, gene combinations associated with altered brain regions were then entered into an enrichment analysis.

**Results:** We found an increased differentiation of connectivity profiles between unimodal and transmodal networks in NDfE, which was particularly pronounced in the patients with persistent seizures at 12-months follow-up (n=10). Differences corresponded to gradient score reductions in a visual network and increases in limbic and default mode systems which subserve higher-level cognition. Cortical difference maps were spatially correlated with regional expression of a weighted gene combination. These genes were enriched for disease and ontology terms and pathways previously associated with epilepsy and seizure susceptibility.

**Interpretations:** Large-scale functional hierarchy may be altered from in focal epilepsy from diagnosis and correlate with response to treatment. Combining functional neuroimaging and transcriptional data analysis may provide a framework for understanding the wide-ranging impairments associated with the disorder and mechanistic insight into how gene processes may drive alterations in brain function mediating the genetic risk of epilepsy.

## 1 Introduction

Epilepsy is a debilitating neurological condition characterised by unprovoked recurrent seizures effecting around 1% of the population.^1^ Developments in both neuroimaging and genetic research has provided insight into underlying pathological mechanisms.^2,3^ Despite significant efforts, however, progress in managing epilepsy has been slow, with a third of patients unresponsive to available medication.^4^ There is a need for reliable neural markers to optimise care pathways from the onset of the condition, particularly for focal epilepsies which have been most commonly associated with pharmacoresistance.^4^ Examining patients with a newly-diagnosed focal epilepsy (NDfE) with advanced neuroimaging methods is still rare,^5^ though key to understanding pathophysiological processes independent of effects related to prolonged exposure to anti-seizure medication (ASM) and disease or seizure load.

There is growing consensus that epilepsy is a network disorder, with alterations observed in how discrete brain regions functionally and structurally related to one another.^6,7^ Resting-state functional MRI (rs-fMRI) provides a window into spontaneous co-fluctuations of brain activity between regions. Functional network alterations such as increased hubness^8^ and local connectivity strength^9^ in networks including the deault mode network (DMN) have been identified in focal epilepsy.^10,11^ However, despite some promising whole-brain neural circuitry (the connectome) findings, clear and reproducible results remain elusive and do not fully account for the functional impairments identified in focal epilepsy from diagnosis in domains ranging from low-level sensorimotor performance to higher-order cognitive processes.^12,13^

Recent work has examined connectomes derived from rs-fMRI with gradient mapping.^14^ Gradients represent smooth transitions of connectivity structure along the cortical surface, revealing patterns of large-scale network hierarchy. The principle functional gradient that explains the most variance of brain-wide connectivity captures a transition from unimodal sensorimotor areas to transmodal networks including the DMN associated with higher-order processing.^15,16^ Gradient alterations have been identified in a number of clinical populations including autism^17^ and generalised epilepsy.^18^ Compared to traditional edge-based connectivity, low-dimensional connectome representations show increased phenotypic prediction accuracy across a range of cognitive domains,^19^ suggesting potential clinical utility for diagnosing and monitoring neurological conditions.

Taking advantage of recent methodelogical advances that allow imaging data to be examined alongside transcriptional datasets, large-scale functional networks have also correlate with variation in cortical gene expression.^20^ A study also showed that features of brain structure in patients with psychosis was associated with regional brain expression of schizophrenia-related genes.^21^ Given the growing evidence that genetic factors play a key role in the development of focal epilepsies,^22,23^ combining functional connectomic with transcriptional data may provide insights into the pathophysiology of epilepsy and clues that lead to novel treatment targets.

The aim of this case-control study was to examine rs-fMRI gradient mapping in conjunction with transcriptional data to: (1) identify alterations in functional network architecture in patients with NDfE; and (2) examine whether such alterations are associated brain expression of epilepsy-related genes. We expected to find principle gradient score differences between patients (particularly those with poorer seizure outcomes) and controls and that affected regions would be associated with an over-representation of gene patterns/pathways previously associated with epilepsy and seizure susceptibility.

## 2 Materials and methods

### 2.1 Participants

We analysed data from 27 patients (12 females) with NDfE (mean age, 33.1±11.3, range, 18-57) recruited over a 2-year period from clinics at the Walton Centre NHS Foundation Trust, Liverpool, UK. Focal epilepsy was diagnosed by epileptologists based on the assessment of seizure semiology in accordance with International League Against Epilepsy (ILAE) operational classifications.^24^ Patients with known progressive neurological disease or provoked, acute symptomatic, unclassified, or primary generalized seizures were not recruited. To increase sample size, we did not limit recruitment to drug-naïve patients. In-depth clinical information on these patients has been previously published.^11^ Scanning took place an average of 3.7 months (±2.9, range, 1–11 months) after diagnosis – a time interval when we did not expect adverse medication-related effects on brain function. All patients were followed up one year after scanning to determine current seizure status and response to anti-epileptic medication. All patients were followed up one year after scanning to determine current seizure status and response to anti-epileptic medication treatment. We also scanned 36 neurotypical controls (22 females) with no neurological or psychiatric history, matched to patients for age (mean age 33.7±11.6, range 18-58). Written, informed consent for this study was obtained from all participants in accordance with the Declaration of Helsinki and board which provided full ethical approval (North West - Liverpool Central Research Ethics Committee).

### 2.2 MRI acquisition

All patient and control participants were scanned at the Liverpool Magnetic Resonance Imaging Centre (LiMRIC), University of Liverpool. We used a 3T MR system (Siemens Trio) to acquired 3D T1□weighted (T1w) and resting□state functional MRI (rs-fMRI) data. A Magnetization Prepared Rapid Gradient Echo (MPRAGE) sequence was used for T1w images (TE = 5.57 ms; TR = 2040 ms; TI = 1,100 ms; slice thickness = 1 mm; voxel size = 1 mm × 1 mm; 176 slices; flip angle = 8) and the rs-fMRI consisted of a 6 minute T2□weighted sequence (turbo spin echo with variable flip angle; TE = 355 ms; TR = 3,000 ms; slice thickness = 1 mm; voxel size = 1 mm x 1 mm; Turbo factor = 209).

### 2.3 Imaging analysis

We preprocessed rs-fMRI data using SPM12 (Welcome Trust Centre for Neuroimaging, University College London, United Kingdom; http://www.fil.ion.ucl.ac.uk/spm/) in a Matlab v.9.0 framework (The Mathworks Inc, USA). In brief, images were realigned, slice□time corrected, spatially normalized to the Montreal Neurological Institute (MNI) space and spatially smoothed with an 8□mm full□width half□maximum Gaussian kernel. Spatially preprocessed rs-fMRI data were analysed using the Functional Connectivity Toolbox (CONN).^25^ Data were temporally filtered with a band-pass filtering (0.008–0.09 Hz) to reduce low□frequency drift and noise effects. After co-registering functional and T1w data in MNI152 space through a combination of linear and non-linear transformations, MRI quality and preprocessing were visually and quantitatively evaluated. A component□based noise correction approach was implemented to remove variance related to confounding signals including movement in the scanner and blood□oxygen□level□dependent (BOLD) response originating in white matter and ventricles using CONN’s default denoising pipeline (CompCor).^26^ No participants had to be removed from the analysis due to low-quality structural or functional MRI or excessive head motion in rs-fMRI data (translation > 3 mm, rotation > 1°).

The mean functional time-series from 400 regions, based on the 400 Schaefer parcellation,^27^ were extracted for each individual. A 400 x 400 functional brain connectivity matrix was generated for each participant by computing bivariate correlations between each pair of regions with coefficients transformed into z-scores (Fisher z).

### 2.4 Connectome gradient analysis

Fig. 1 shows an outline of the analysis pipeline implemented in this study. We extracted gradients from the functional connectivity matrix of each participant using the BrainSpace Toolbox.^28^ The matrix was thresholded with a sparsity parameter of 90 and similarity of connectivity profile between regions was computed with a cosine similarity kernel, following prior approaches.^14^ Ten gradient components with decreasing explained variance were computed using diffusion map embedding, a nonlinear dimension reduction approach.^29^ In gradient space, regions with similar patterns of functional connectivity are closer to one another than regions with different functional connectivity profiles. The alpha (α) parameter which controls the influence of sampling density was set to 0.5 to retain global gradient space relations between regions. To allow between-individual comparisons, Procrustes rotation was used to align gradient components of each participant to a gradient template, computed from the average connectivity matrix of all participants.^30^

**Fig. 1.**
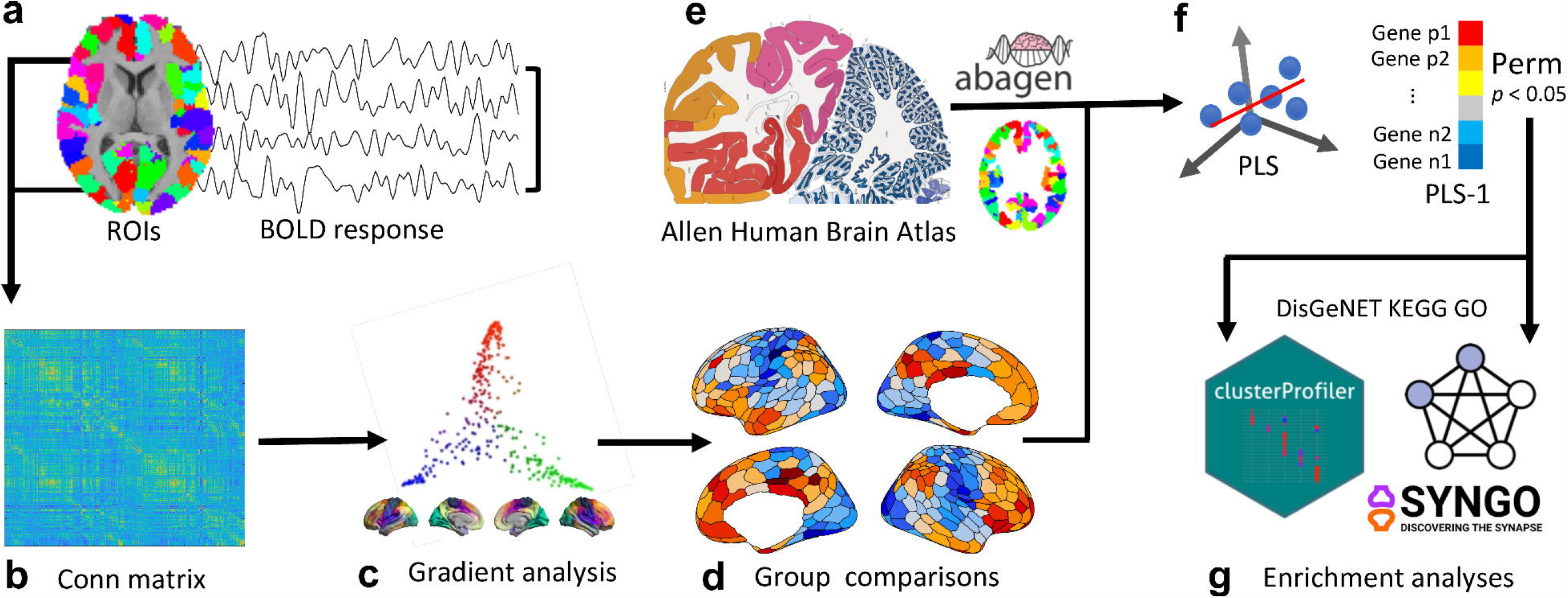
Study analysis pipeline. (a) Preprocessed resting-state fMRI (rs-fMRI) timeseries for each region of interest (ROI) pair were correlated (b) forming a functional connectivity (conn) matrix for each participant (c) and used as input for a gradient analysis. (d) Following group comparisons of principle gradient scores spatial case-control t statistics were used to predict (e) regional gene expression (Allen Human Brain Atlas) for each ROI in (f) a PLS regression analysis with permutation testing. Genes with Pperm < 0.05 were then examined in (g) functional gene enrichment analyses.

We focused on the first/principle gradient with the best defined functional implications, as shown in previous work.^17,31^ To compare gradient scores mapped back onto the cortical surface in NDfE and control groups, and in PS and SF patient subgroups, we used general linear models (GLM) that controlled for the effects of age and sex. PALM, a non-parametric permutation-based FSL tool,^32^ was used for family-wise error rate (FWER) correction of p-values (10,000 permutations). We also tested for group differences within seven canonical functional networks with PALM (10,000 permutations), correcting for age and sex.

### 2.5 Gene expression analysis

We ran a transcriptomic analysis of case–control gradient differences using the AHBA, a transcriptomic dataset with gene expression measurements collected from six post mortem adult brains aged between 24 and 57 years (human.brain-map.org).^33^ AHBA data were collapsed into Schaefer atlas regions and combined across donors using differential stability with the abagen python toolbox (https://github.com/rmarkello/abagen), resulting in a 400 × 15632 regional transcription matrix. Following a previously outlined procedure,^21^ we used gene expression levels to predict case-control t-statistics for the principal gradient (the response variable) with a partial least squares (PLS) analysis. The first component of the PLS (PLS-1) consists of the linear combination of weighted gene expression values most strongly related to the cortical map of case–control differences.^21^ Statistical significance of the weight associated with each gene was determined by permuting the response variables 10,000 times (resampling with replacement of the 400 cortical regions). Gene weights that fell below the 0.025 percentile and above the 0.975 percentile of the null distribution were classified as significant contributors to PLS-1 at an alpha of 0.05, and selected for further analysis (Pperm < 0.05).

We used Search Tool for the Retrieval of Interacting Genes Database (STRING v11) (https://www.string-db.org/) for the visualisation of protein–protein interaction (PPI) using the STRINGdb R package.^34^ Enrichment of associated functions and pathways was examined by using the clusterProfiler R package.^35,36^ This included disease enrichment analysis based on the DisGeNET (http://www.disgenet.org/), Kyoto Encyclopaedia of Genes and Genomes (KEGG) and Gene Ontology (GO) enrichment analysis with sub-ontologies (Biological Process - BP, Molecular Function - MF, and Cellular Component - CC). Signed PLS-1 weights were used to rank the input set of genes with Pperm < 0.05. We used default setting which included a p-value cutoff of 0.05 with adjustment,^37^ a q-value cutoff of 0.2, a background gene list made up of all genes included in the database, and a minimum and maximum gene set size of 10-500. For more specific insight into synapse function associated with PLS1-derived gene sets, we also explored synaptic enrichment in using SynGO (www.syngoportal.org) with default settings including a background of brain expressed genes.

## 3 Results

### 3.1 Gradient results

The first functional gradient explained 14.3% of connectome variance in rs-fMRI data and consisted of a continuum ranging from low-level sensory systems to transmodal networks, with intermediary networks positioned in between (Fig. 2a). Subgroup analyses confirmed that the variance explained was similar in NDfE and HC groups (t = -0.1, p = 0.9). We compared participant gradient scores using surface-based GLMs controlling for age and sex. Globally, we found increased functional separation between unimodal and transmodal networks in NDfE, which was particularly pronounced in the PS group (Fig 2). Differences corresponded to gradient score reductions compared to controls in the visual network, and increases at the DMN end, with decreased density in the middle of the gradient continuum (Fig. 2e). Brain regions with highest principle gradient values in controls tended to show the greatest increases in patients, and conversely, areas with the lowest negative values in healthy controls had the greatest decrease in patients with NDfE. In other words, differences in regional connectivity profiles were maximal at the unimodal and transmodal anchors of the principal gradient, both in the whole NDfE group (r = 0.73, p < 0.001), and in the PS subgroup (r = 0.67 p < 0.001) compared to controls (Fig. 2c). Group comparisons using non-parametric permutation tests (PALM) revealed increased gradient scores located in prefrontal, temporal and posterior cingulate regions in patient with NDfE compared to HC with reductions in occipital, occipito-temporal and central sensorimotor regions. Compared to the SF group, scores were similarly increased in a posterior cingulate region in patients with PS compared to those that became SF with reductions located in a visual region. We also summarised surface-wide findings according to the seven functional system decomposition^38^ which is part of the Schaefer 400 parcellation scheme (Fig. 2d & 3e). Permutation-based testing (PALM) showed a significant reduction in the visual (t = -2.07, p = 0.023, pCor = 0.12) and sensorimotor (t = -1.78, p=0.042, pCor = 0.224) networks and increases in the limbic network (t = 2.5, p = 0.004, pCor = 0.042) in patient with NDfE compared to HC, though only the limbic network survived FWER correction for comparisons across the seven networks (see pCor). Gradient score increases were found in the DMN in PS compared to SF patients (t = 2.98, p = 0.002, pCor = 0.019).

**Fig. 2.**
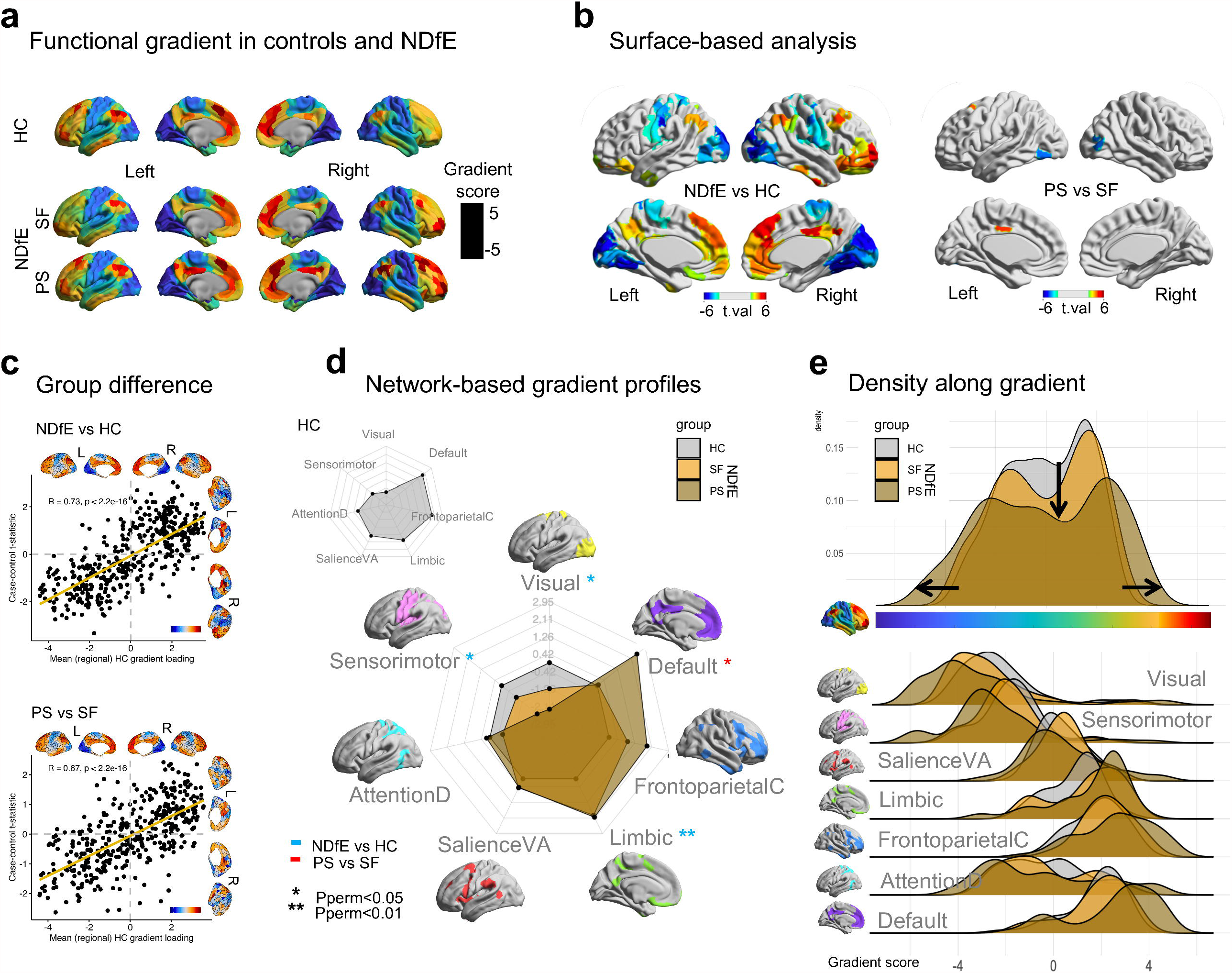
Principle gradient group comparisons. (a) Principle gradient scores shown on the brain surface averaged across healthy controls (HC) and patients with newly diagnosed focal epilepsy (NDfE) subdivided into patients that became seizure free (SF) and those with persistent seizures (PS) in the 12 months after testing. This gradient corresponds to a continuum ranging from unimodal (sensory, motor) regions (dark blue) to transmodal (default mode network - DMN) regions (dark red), with colour similarity depicting similarity in connectivity patterning. While the gradient is visually similar across groups, higher scores can be seen in the DMN core in NDfE, particularly in PS patients. (b) Surface-based statistical comparisons between NDfE and HC and between PS and SF patients, with increases/decreases (t statistics) shown in red/blue for regional with p < 0.05 (FWER correction) in a permutation test (PALM - 10000 permutations). (c) Scatterplot of mean HC principle gradient scores (x axis) vs. case–control t statistic of gradient scores (y axis) for NDfE-HC and PS-SF with associated brain surface representations and correlation values. (d) Network-based permutation testing (PALM) of median gradient scores of patient groups with respect to controls shows significant reductions in visual and sensorimotor systems and increases in limbic and DM networks primarily for PS in spider plots. (e) Global gradient density plot shows greater differentiation in between unimodal and transmodal systems (more extreme gradient scores) with suppressed density in the middle of the gradient in NDfE (particularly PS) compared to HC. Joy plots (below) also show gradient density along the principal gradient by network for HC and NDfE groups (SF and PS).

### 3.2 Gene expression related to gradient difference

We used a PLS regression to examine gene expression patterns correlated with cortical maps of case–control differences for NDfE-HC and PS-SF comparisons. The first component (PLS-1) explained 12% of the variance in the data with associated weights positively correlated with cortical NDfE-HC t-statistics (r = 0.58, p < 0.001) (Fig. 3a). In predicting cortical PS-SF t-statistics, the first component explained 11.5% of variance with weights also positively correlated with the predictor (r = 0.41, p < 0.001). We identified 3,258 genes at the chosen permutation-derived p-value threshold (Pperm < 0.05). These positive correlations indicate that positively weighted genes are over-expressed in cortical regions where principle gradient scores increased for patients with NDfE and PS, while genes with negative weighting are over-expressed for these groups in regions with decreased gradient scores (Fig. 3c).

**Fig. 3.**
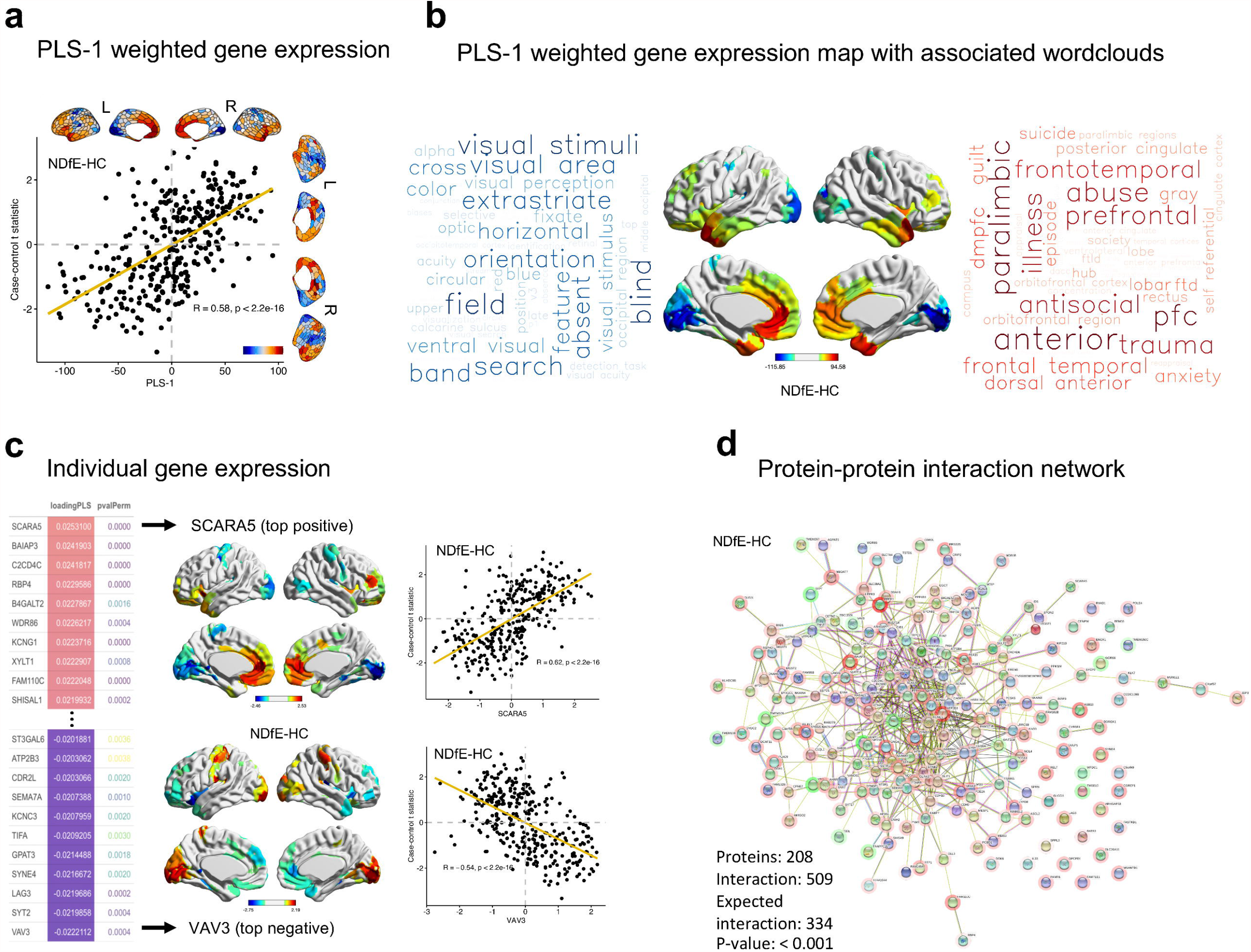
Gene expression profiles related to differences between patients with newly diagnosed focal epilepsy (NDfE) and healthy controls (HC) in principal gradient scores. (a) Scatterplot of regional PLS-1 scores (weighted sum of 15,632 gene expression scores) (x axis) vs. case-control differences (NDfE–HC t statistics) in regional gradient scores (y axis) with associated brain surface representations and correlation values. (b) Cortical map of regional PLS-1 scores with separate wordclouds for negative (blue) and positive scores generated through a NeuroQuery image decoding meta-analysis. (c) The 10 strongest positively and negatively weighted genes on PLS-1 for the NDfE-HC comparison with associated permutation-derived p-values (pvalPerm) (left), alongside examplar brain surface images and scatterplots (case-control t statistics vs. regional gene expression) for the top positively rated gene (SCARA5) and the top negatively rated gene (VAV3) (right). (d) The protein-protein interaction network (PPI) for significant (Pperm < 0.01) PLS-1 derived genes from the NDfE-HC comparison. Coloured nodes indicate query proteins and first shell interactors (as per the STRING default settings) and negatively/positively weighted genes are shown with a green/red halo. Clustered genes are likely to have similar functions. The p-value represents the probability of an equal or greater number of interactions occurring by chance.

### 3.3 Enrichment analysis of genes related to gradient difference

We found 943 genes with Pperm < 0.05 for the NDfE-HC comparison (263 negatively weighted) (see Dataset1). The protein-protein interaction network for PLS-derived genes from the NDfE-HC comparison had 208 connected proteins and 509 edges/interactions, significantly more than the 334 edges expected by chance (Pperm < 0.001) (Fig. 3d). We also tested the ranked PLS-derived gene sets for significant disease, GO and KEGG pathway enrichment (Dataset2). Amongst over-represented genes for the NDfE-HC comparison, eight out of the 17 significant (p.adjusted BH-FDR method < 0.05) disease enriched terms (DisGeNET) referred to forms of epilepsy or seizures including focal (geneRatio = 22/829; p.adjust = 0.024), tonic-clonic (geneRatio = 28/829; p.adjust = 0.022) and absence (geneRatio = 23/829; p.adjust = 0.008) seizures (Fig. 4a) while other terms related to impulsive behaviour (geneRatio = 25/829; p.adjust = 0.035), melatonin deficiency (geneRatio = 8/829; p.adjust < 0.001) and pain-related conditions such as hyperalgesia (see Fig. 4). Genes implicated in epilepsy-related disease networks (Fig 5a) were among those that formed the most strongly interconnected cluster of nodes in the PPI network (e.g., KCNC1, SCN1B and GABRA5) (Fig. 3d). The KEGG pathway analysis found significant enrichment in a number of cell signalling pathways including “neuroactive ligand-receptor interaction” (geneRatio = 26/338; p.adjust = 0.048) and “calcium signaling pathway” (geneRatio = 20/338; p.adjust = 0.048), as well as glutamatergic-related mechanisms such as “glutamatergic synapse” (geneRatio = 12/338; p.adjust = 0.048) and “glutamatergic metabolism” (geneRatio = 8/338; p.adjust = 0.048). The highest significant gene ratios in GO sub-ontologies included “cellular response to metal ion” (BP: geneRatio = 30/869; p.adjust < 0.001), “synaptic membrane” (CC: geneRatio = 48/906; p.adjust < 0.001), and “metal ion transmembrane transporter activity” (MF: geneRatio = 47/897; p.adjust < 0.001). Results of the enrichment analysis are provided in Dataset2. The SynGO analysis found more presynaptic-related genes (CC:“presynapse”, geneRatio = 68/536, -log10 q-value > 8; BP:“process in the presynapse”, geneRatio = 38/269, -log10 q-value > 5) than postsynaptic genes (CC:“postsynapse”, geneRatio = 58/624, -log10 q-value > 3; BP:“process in the postsynapse”, gene count = 28/218, -log10 q-value > 4) (Fig. 4b).

**Fig. 4.**
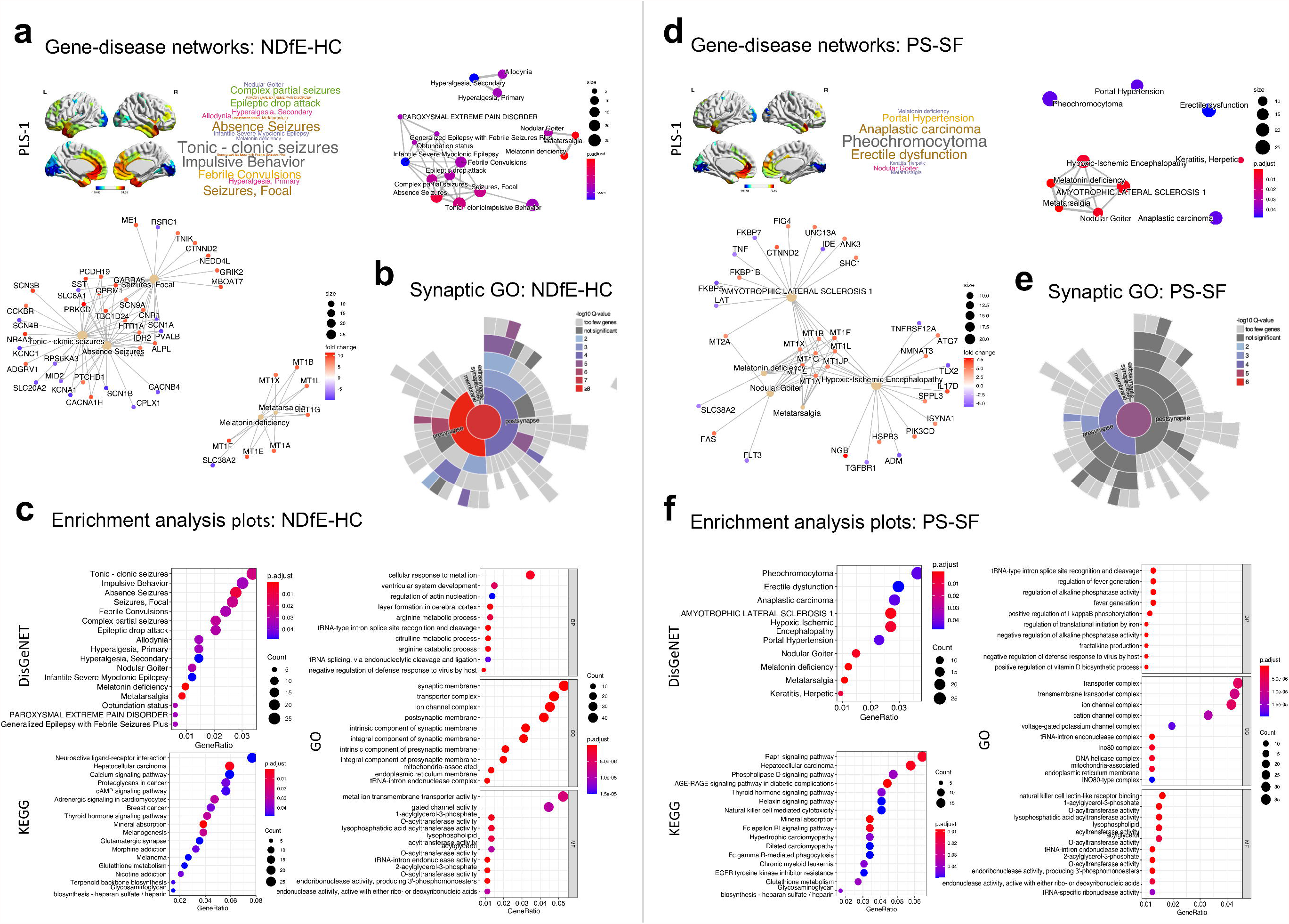
Enrichment analysis of genes transcriptionally related to case-control differences in principle gradient scores (PLS-1). (a) Brain surface maps for PLS-1 (weighted sum of 15,632 gene expression scores) are shown (left) with significantly enriched terms derived from a disease enrichment analysis (DisGeNET) (right) comparing gradient differences in patients with newly diagnosed focal epilepsy (NDfE) and healthy controls (HC). Analysis results are presented as gene-disease (below) and disease-disease networks (right) where edges are weighted by the ratio of overlapping gene sets which cluster together and indicate functional similarity. (b) Sunburst plots for synaptic gene ontology (SynGO) analysis results which show median significance values (-log 10 Q-value) for SynGO-annotated cellular components, as determined by Wilcoxon rank-sum test followed by Bonferroni correction at α 0.05. (c) Bubble plots for enrichment analyses including gene-disease and variant-disease associations (DisGeNET) (top left) Kyoto Encyclopaedia of Genes and Genomes (KEGG) pathways (bottom left) and Gene Ontology (GO) separated into sub-ontologies (Biological Process - BP, Molecular Function - MF, and Cellular Component - CC) (right). Terms are ordered by gene ratio displaying count (bubble size) and enrichment score as adjusted p values. (d, e, f) The equivalent enrichment analysis plots comparing patients that continued to have persistent seizures (PS) with those that became seizure free (SF) in the 12 months after testing are shown on the right.

We found 838 genes (282 negatively weighted) for the PS-SF comparison (Dataset1). Notably, no significant disease enriched terms related to epilepsy or seizure were found in this comparison. The significant term with the highest gene ratio was Pheochromocytoma (geneRatio = 27/740; p.adjust < 0.044) while the most significant term was melatonin deficiency (geneRatio = 9/829; p.adjust < 0.001). The significant terms with the highest gene ratio for the PS-SF comparison were, KEGG: “Rap 1 signaling pathway”, geneRatio = 19/294, p.adjust < 0.01; GO:BP: “tRNA-type intron splice site recognition and cleavage”, geneRatio = 10/788, p.adjust < 0.001; GO:CC: “transporter complex”, geneRatio = 36/818, p.adjust < 0.001; GO:MF: “natural killer cell lectin-like receptor binding”, geneRatio = 13/810, p.adjust < 0.001. The SynGO analysis for PS-SF also found more presynaptic-related genes (CC: “presynapse”, geneRatio = 49/536, -log10 q-value > 4; BP: “process in the presynapse”, geneRatio = 28/269, -log10 q-value > 2) than postsynaptic genes (CC: “postsynapse”, geneRatio = 45/624, -log10 q-value = ns; BP: “process in the presynapse”, geneRatio = 23/218, -log10 q-value > 2) (Fig. 4e). Results for the enrichment analyses are presented in full in Dataset2.

## 4 Discussion

In this study, we examined whether alterations in large-scale functional networks and the potential genetic basis of such alterations can be identified in patients with NDfE and whether they are associated with future response to treatment (i.e., seizure status) using gradient mapping of rs-fMRI. There is evidence that hierarchy in functional systems such as sensory, motor, and default mode networks is a key principle of brain organisation that determines how information flows across the cortex.^39,40^ Gradient mapping linked to what is known about the distribution of gene expression across the brain can offer a more global perspective on network alterations in focal epilepsy with potential to account for the wide-ranging sensory, motor and higher-order cognitive impairments associated with the condition from diagnosis.

Our gradient mapping results showed that the component explaining the most variance in the connectivity data across participants corresponded to a dimension spanning from low-level sensory to transmodal (default mode) systems with other networks in-between [Fig. 2a], in line with previous work.^39^ Crucially, we found increased functional separation between sub-systems in patients with NDfE compared to controls. In our data, this was characterised by lower gradient scores in unimodal regions (visual and rolandic cortices) and higher scores in transmodal hubs (PCC, lateral and medial PFC), with reduced density in mid-gradient networks, such as attentional systems (Fig. 2). These differences were all more pronounced in patients that continued to experience seizures in the following 12 months (PS) after scanning. The general direction of these findings is in line with a recent study that also found principle gradient expansion in patients with genetic generalised epilepsy,^18^ in notable contrast to the global contraction of unimodal and transmodal networks observed in patients with ASD.^17^ Together, clinical gradient findings provisionally suggest that global expansion may be a broader phenotype of epilepsy or seizure-related brain architecture distinguishable from other brain disorders.

In specific reference to the transmodal end of the principle gradient, there is evidence from rs-fMRI and EEG–fMRI studies that DMN connectivity is altered both in focal epilepsy.^10,41^ It is widely acknowledged that DMN core nodes play a key role in internally-focused cognition as well as aspects of language processing and memory,^42^ neuropsychological domains also known to be impaired in patients with focal epilepsy.^13,43^ In our findings, gradient score difference specifically in the DMN was driven by the PS group (Fig. 2d), indicating that this may represent a promising early marker of pharmacoresistance and treatment outcome. It also leaves open the possibility that response to medication and illness duration could partly contribute to DMN abnormalities identified in previous studies,^44,45^ given that these have tended to examine patients with refractory epilepsy. There is also some evidence of dysfunction in networks involved in motor and sensory processes which have also been associated with interictal perceptual as well as cognitive deficits.^46–48^ Our results suggest that diverse neural and behavioural phenotypes of patients with NDfE could be explained in a more unified framework by alterations in a specific large-scale mode of connectivity, one that determines how information flows from sensory and motor to DMN systems through salience, limbic, frontoparietal and attention-related networks. Future studies using neuropsychological tests to directly examine the relationship between cognitive performance and functional gradients are advocated and in progress.^49^ Intrinsic connectivity gradients have recently been used to capture task-related functional reorganisation of episodic memory processes, as well as language and working memory systems in temporal lobe epilepsy.^50,51^

To link our gradient findings of stronger differentiation between unimodal and transmodal systems in NDfE and PS to the histologically measured distribution of gene expression in the brain, we first used a PLS regression to identify weighted gene combinations with cortical expression maps that best matched maps of principal gradient differences between groups (PLS-1).^21^ We then ran a series of enrichment analyses to examine diseases and functional processes that have been associated with identified gene combinations in previous genetics and functional genomics research. We found strong correlations between case-control differences of principle gradient scores and cortical gene expression maps for group comparisons (PLS-1). This tight coupling suggests that there is a genetic basis to functional connectome differences between clinical and non-clinical populations and is supported by prior work which found a correlation between large-scale functional networks and variation in cortical gene expression.^20^

Comparing patients with NDfE with controls, we found that nearly half of all significant disease enriched terms (8/17) referred to epilepsy or seizure subtypes, indicating that genes previously associated with these terms were over-represented in the weighted gene combinations related to between-group differences in principle gradient scores. Additional investigation revealed that proteins coded by the PLS-derived genes formed a dense PPI cluster, with genes significantly enriched for a number of KEGG pathways and GO processes associated with abnormal neuronal function in epilepsy. For example, we found high enrichment scores in a number of cell signalling pathways known to enhance susceptibility to epileptic seizures such as neuroactive ligand-receptor interaction,^52^ glutamatergic processes implicated in aberrant neuronal excitability,^53^ pathways involving ion dynamics and features of synaptic transmission,^54^ and calcium signaling pathways liked to neuronal synchronization and hyperexcitability.^55^

In contrast, no terms specified forms of epilepsy or seizures in the PS-SF group comparison. This makes sense given that this comparison was between patients with gene expression profiles consistent with a diagnosis of epilepsy. Enriched disease terms instead referred to conditions including melatonin deficiency and tumor variants, that together may indicate a higher predisposition for a neuroendocrine/autonomic imbalance with some previous evidence of indirect links to seizures susceptibility.^56,57^ A number of membrane transporter processes that mediate drug uptake and efflux were also identified,^58^ as well as pathways that regulate intercellular signaling (e.g., Phospholipase D), particularly in cells that are under stress conditions.^59^ Significant terms highlighted here could provide a starting point for a targeted, larger scale exploration into response to anti-seizure medication which examines patterns of inter-related diseases, pathways and processes associated with functional and structural brain connectivity hierarchy.

Although the use of a newly diagnosed cohort is a major strength of this study, the sample size was relatively small, particularly for the patient outcome comparison (PS-SF). There are also inherent limitations associated with relying upon gene expression data derived from a different sample (AHBA), though useful brain-specific insights have been gleaned using this approach,^21^ which has the advantage of being more easily applied to clinical settings. The replication of findings in a larger sample with associated genome-wide whole blood transcriptome profiling and neuropsychological data will be useful in determining the diagnostic and prognostic potential of combining gradient mapping with gene expression for focal epilepsy.

In conclusion, the findings of this study suggest that large-scale functional networks in rs-fMRI are altered from early stages of focal epilepsy, and correlate with response to treatment. The approach offers a global perspective on network alterations in focal epilepsy with potential to provide a unifying account for the previously characterized wide-ranging sensory, motor and higher-order cognitive impairments associated with the condition from diagnosis. When combined with transcriptional profiling, such alterations can highlight pathways and biological processes relevant to epilepsy and pharmacoresistance. This multi-method approach can contribute to bridging the gap between macro- and micro-scale mechanisms in neuroscience and provide a framework for understanding the interaction between brain processes with the aim of deep phenotyping that leads to precision medicine in the treatment of epilepsy.

## Data Availability

Data are available from the authors upon reasonable request.

## 5 Acknowledgements

The authors thank patients and controls for their time and participation and the UK Medical Research Council for financial support (grants MR/S00355X/1 and MR/K023152/1 awarded to SSK).

## 7 Author contributions

CEdB was involved in the conception and design of the study, analysis of data, and manuscript drafting; SSK, AGM, LC and BCB contributed to the design and conception of study and text drafting; BA contributed to acquisition of data.

